# Trends in Cardiac Arrest Among Heart Failure Patients Aged 25 and Older in the United States: Insights from the CDC WONDER Database

**DOI:** 10.1101/2024.12.04.24318529

**Authors:** Abdullah Naveed Muhammad, Sivaram Neppala, Himaja Dutt Chigurupati, Ahila Ali, Muhammad Omer Rehan, Sowjanya Kapaganti, Rabia Iqbal, Hamza Naveed, Mushood Ahmed, Gregg C. Fonarow, Sourbha Dani

**Author notes:** Corresponding Author: Sivaram Neppala, MD, Assistant Professor, Department of Medicine, University of Texas Health Science Center, San Antonio, TX.

## Abstract

**Background:** Cardiac arrest (CA) is a significant cause of mortality in patients with heart failure (HF). Despite improvements in care, disparities in CA-related mortality among HF patients persist. This study seeks to explore demographic patterns and mortality rates.

**Methods:** CDC WONDER (1999-2023) data was analyzed to assess CA-related mortality in HF patients aged≥25. Using the Joinpoint regression analysis, we calculated age-adjusted mortality rates (AAMR) per 100,000 patients and average annual percentage changes (AAPC) to analyze the mortality trends.

**Results:** CA is responsible for a staggering 1,139,963 deaths among HF patients. The AAMRs have decreased from 25.3 in 1999 to 20.6 in 2023 (AAPC: -0.88, p-value < 0.01), the decline was most significant from 1999 to 2011 (APC: -2.95, p < 0.01) before a concerning rise from 2018 to 2021 (APC: 5.10, p-value < 0.01). Disparities are evident, with men facing higher AAMRs than women (24.6 vs. 17.6) and Black individuals at the highest risk (AAMR: 28.8) followed by Hispanics (AAMR: 22.8). Regionally, Mississippi reports the highest AAMR at 52.5, contrasting with Maryland’s lowest rate of 4.3. Moreover, rural areas exhibited higher AAMRs than urban settings (20.9 vs. 20.1).

**Conclusion:** The rising mortality rates from CA in HF patients—especially among men, Black individuals, and those in rural areas—require our immediate attention. By implementing targeted interventions and enhancing access to healthcare, we can reduce these disparities and improve patient outcomes.

## Introduction

Heart failure (HF) is a chronic and debilitating condition that represents a major global health crisis, impacting the lives of over 64 million adults worldwide as of 2017 (1). In the U.S., about 6 million adults aged 20 and older have HF, a number expected to reach 8 million by 2030 (2). Despite advances in treatments, HF patients have a much higher risk of cardiac arrest (CA), which accounts for 30-50% of deaths in these patients. Those with systolic dysfunction are 6 to 9 times more likely to experience sudden cardiac death compared to the general population (3,4). The link between CA and HF often involves abnormal heart rhythms and changes in heart structure (5,6). Understanding these trends is crucial for improving this vulnerable group’s risk assessment and preventive strategies.

Research shows that younger adults with inherited heart diseases and older adults with ischemic or non-ischemic cardiomyopathy are at greater risk (7-9). Notably, disparities in health outcomes related to race, income, and location hinder access to care for cardiac arrest patients with heart failure. For example, Black individuals with heart failure experience higher mortality rates than White individuals, often due to differences in genetics, healthcare access, and treatment adherence (10-12). Additionally, women with heart failure generally fare better than men (13). These disparities highlight the urgent need for targeted initiatives and equitable healthcare resources to reduce cardiac arrest risk and improve outcomes for all heart failure patients.

Recent research shows that new HF treatments, such as following medical guidelines and using implantable cardioverter-defibrillators (ICDs), have greatly improved patient survival rates (14, 15). However, many challenges remain, including differences in income, access to healthcare, and the lack of effective treatments among vulnerable groups (16, 17). Additionally, younger adults with HF, especially those with non-ischemic causes, are often left out of clinical trials.

### This leads to gaps in prevention and overall care for these patients

This study explores changing trends and differences in outcomes related to CA in people with HF. It focuses on how clinical risk factors and demographic inequalities interact. The goal is to find practical ways to improve prevention strategies, assess risks, and enhance healthcare for this vulnerable population.

## Methods

### Study design

The data utilized in this study was procured from the Centers for Disease Control and Prevention-Wide-ranging ONline Data for Epidemiologic Research (CDC-WONDER) database, a trusted and comprehensive source of death certificates. Covering the period from 1999 to 2023, the dataset specifically focused on fatalities attributed to cardiac arrest in individuals with heart failure, employing ICD-10 codes for accuracy. This extensive dataset incorporates death certificates from all 50 states and the District of Columbia and has previously facilitated the analysis of mortality trends across various diseases, targeting adults aged 25 years and older.

This investigation relied on de-identified, publicly available datasets generated by government institutions. While obtaining Institutional Review Board (IRB) approval was not a prerequisite due to the data’s nature, the study proactively conformed to the Strengthening the Reporting of Observational Studies in Epidemiology (STROBE) guidelines, enhancing its findings’ reliability and transparency.

## Study cohort

This study focused on adults aged 25 and older who were diagnosed with heart failure between 1999 and 2023. We rigorously analyzed death records from the Multiple Causes of Death Public Use registry to identify cases of mortality linked to cardiac arrest within this specific population. By employing the International Classification of Diseases (ICD) codes, we specifically targeted instances of cardiac arrest (I46.0, I46.1, I46.9) and heart failure (I11.0, I13.0, I13.2, and I50.x), ensuring a precise and thorough examination of these critical health outcomes.

### Data extraction

This study meticulously compiled a detailed dataset capturing essential mortality-related factors, which include population size, year of death, geographic location, demographic characteristics, urban-rural classification, regional breakdown, and state-specific categorizations. The demographic information encompasses critical variables such as age and race/ethnicity. The location of death is classified into various healthcare environments, including outpatient settings, emergency rooms, and inpatient facilities, as well as determining cases of death on arrival or where the status remains unknown, alongside home, hospice, and nursing homes/long-term care facilities. Race and ethnicity are categorized into Hispanics, non-Hispanic (NH) Whites, NH Blacks, and NH others. Derived from death certificates, these meticulously gathered parameters have been previously validated through the CDC WONDER database analysis, ensuring a comprehensive understanding of mortality trends and their implications.

To effectively gauge the population, we employed the National Center for Health Statistics Urban-Rural Classification Scheme, which categorizes counties as either urban (including large central metropolitan, large fringe metropolitan, medium metropolitan, and small metropolitan) or nonmetropolitan (encompassing micropolitan and noncore areas) based on the 2013 U.S. Census. Leveraging definitions from the U.S. Census Bureau, regions were further delineated into the Northeast, Midwest, South, and West, enhancing the resolution and applicability of our findings. By employing these robust classifications, our study provides crucial insights for understanding mortality patterns across diverse populations.

### Statistical analysis

The study comprehensively analyzed the crude mortality rate and the Age-Adjusted Mortality Rate (AAMR) per 100,000 individuals to investigate nationwide mortality trends thoroughly. This process involved meticulously calculating the total number of fatalities linked to cardiac arrest within the heart failure patient population for each year examined. In line with established research practices, the AAMR was computed by standardizing these cardiac arrest-related deaths against the 2000 U.S. population while generating 95% confidence intervals for accuracy. To derive the annual percent change (APC) and its associated 95% confidence interval in AAMR, we utilized the JoinPoint Regression Program (Joinpoint V 4.9.0.0, National Cancer Institute, Bethesda, MD, USA). Using AAMRs allowed for meaningful comparisons of mortality rates across various populations and historical contexts. By analyzing these rates, the study illuminated mortality trends and highlighted noteworthy fluctuations over time by applying log-linear regression models.

## Results

Between 1999 and 2023, cardiac arrest in individuals with heart failure was responsible for a total of 1,139,963 deaths among adults aged 25 years and older in the United States (Supplementary Table 1). The distribution of these fatalities varied across different healthcare settings, with the majority occurring in medical facilities (49.8%). Additionally, 23.0% of the deaths took place within the decedents’ homes, 22.1% occurred in nursing homes or long-term care facilities, 1.4% transpired in hospice facilities, and 3.3% were reported at other locations (Supplementary Table 2). The central illustration summarizing the study’s characteristics and findings is presented in Fig. 1.

**Fig. 1:**
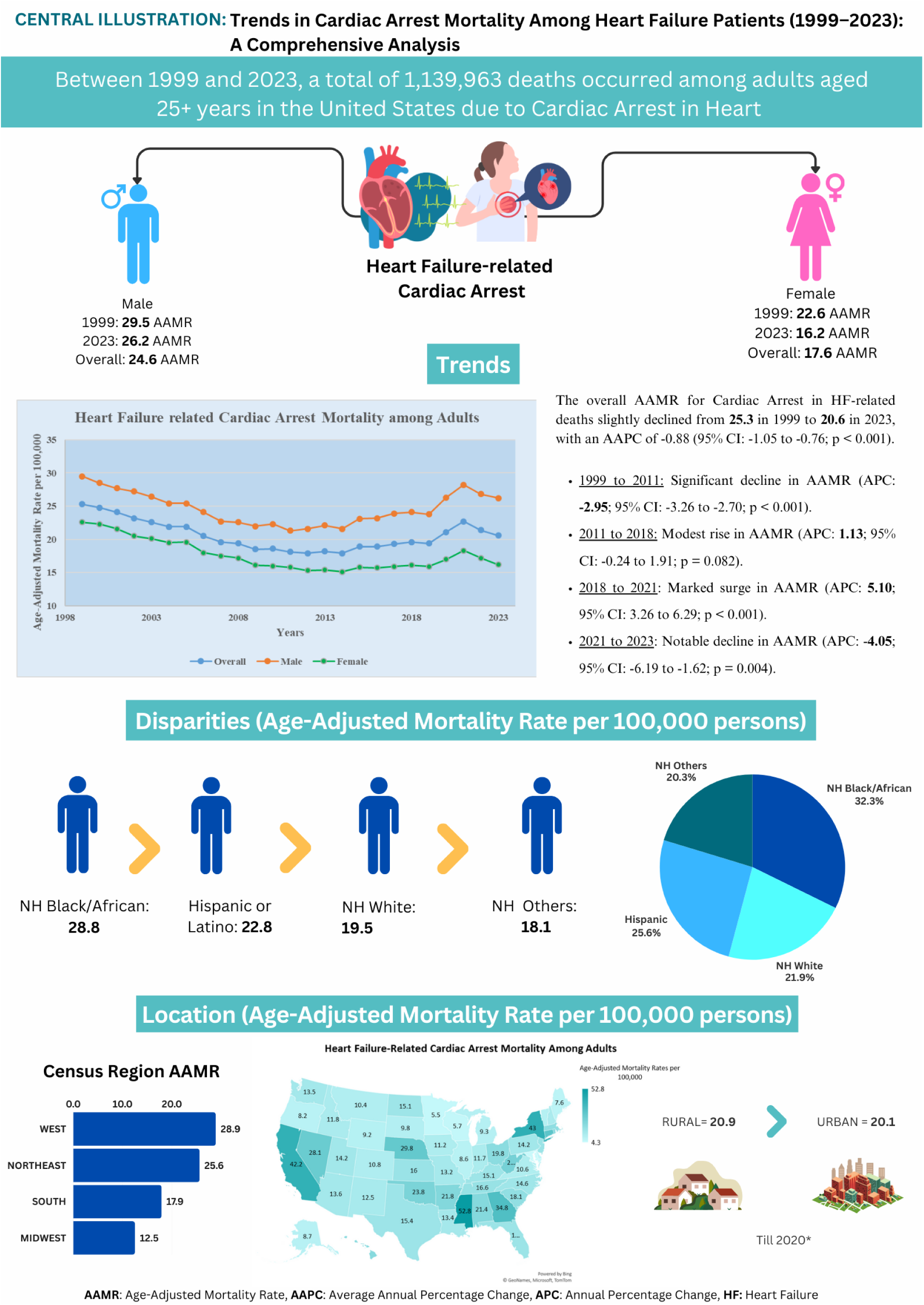
Central Illustration: Trends in Demographics and Disparities in Cardiac Arrest-Related Mortality in Heart Failure Patients in the United States: 1999 to 2023

### Annual Trends for Cardiac Arrest in Heart Failure-Related Age-adjusted Mortality Rate (AAMR)

The age-adjusted mortality rate (AAMR) for CA-related deaths among adults with HF has shown a gradual yet significant decline from 25.3 in 1999 to 20.6 in 2023. This change corresponds to an Average Annual Percentage Change (AAPC) of -0.88 (95% Confidence Interval [CI]: -1.05 to -0.76) (p-value < 0.000001). A particularly significant reduction occurred from 1999 to 2011, marked by an Annual Percentage Change (APC) of -2.95 (95% CI: -3.26 to -2.70) (p-value < 0.000001). However, the period from 2011 to 2018 did not change significantly, with an APC of 1.13 (95% CI: -0.24 to 1.91) (p-value = 0.08). Subsequently, there was a surge in mortality rates from 2018 to 2021, indicated by an APC of 5.10 (95% CI: 3.26 to 6.29) (p-value < 0.000001). Fortunately, this trend reversed from 2021 to 2023, showing a significant decline with an APC of -4.05 (95% CI: -6.19 to -1.62) (p-value = 0.0004) (Supplementary Table 3).

### Cardiac Arrest in Heart Failure-Related AAMR Stratified by Sex

Throughout the study period, it is evident that adult men faced significantly higher AAMRs than adult women, with men reporting an overall AAMR of 24.6 (95% CI: 24.3-25.0) compared to 17.6 for women (95% CI: 17.4-17.8), p <0.0001. Notably, from 1999 to 2023, both genders experienced a decline in AAMR; however, this decrease was markedly more pronounced among women. The AAPC for men was measured at -0.49 (CI: -0.75 to -0.27; p-value = 0.0008), while for women, the AAPC was significantly higher at -1.40 (CI: -1.60 to -1.25; p-value < 0.000001).

For men, the AAMR notably dropped from 29.5 in 1999 to 22.3 in 2010 (APC: -2.83; 95% CI: - 3.66 to -2.30; p-value = 0.01). This was followed by a period from 2011 to 2018 with no significant changes (APC: 0.98; 95% CI: -2.69 to 1.91; p-value = 0.24). However, a dramatic increase was observed from the year 2018, with the AAMR rising to 28.2 by 2021 (APC: 5.82; 95% CI: 0.69 to 7.45; p-value = 0.03), and with no significant difference noted from 2021 to 2023 (APC: -2.46; 95% CI: -5.69 to 2.61; p-value = 0.19).

Similarly, the AAMR for women initially decreased from 22.6 in 1999 to 15.3 in 2012 (APC: - 3.10; 95% CI: -3.54 to -2.84; p-value < 0.000001). This declining trend continued without significant variation until 2018 (APC: 0.78; 95% CI: -2.95 to 1.69; p-value = 0.48). However, by 2021, a striking rise in AAMR was recorded, reaching 18.3 (APC: 4.31; 95% CI: 2.11 to 5.76; p-value = 0.001), leading to a significant decline to 16.2 by 2023 (APC: -4.99; 95% CI: -7.42 to - 1.96; p-value = 0.002) (Supplementary Table 4 and Fig. 2).

**Fig. 2:**
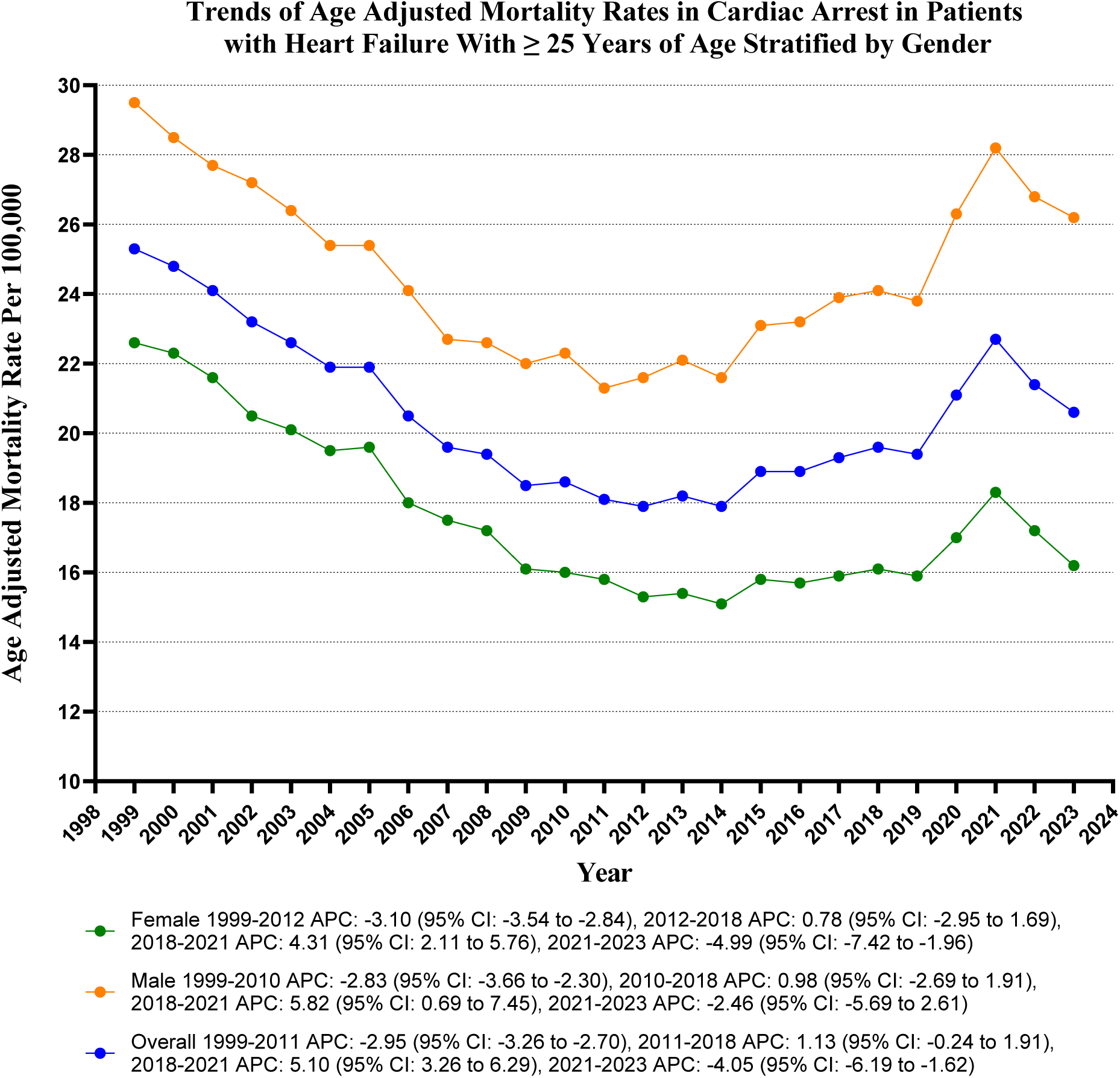
Overall and Sex-Stratified Cardiac arrest related age-adjusted mortality rates per 100,000 Adults with Heart Failure in the United States, 1999 to 2023

### Cardiac Arrest in Heart Failure-Related AAMR Stratified by Race/Ethnicity

Our data revealed that non-Hispanic (NH) Blacks experience the highest AAMRs, followed by Hispanics, non-Hispanic Whites, and non-Hispanic Others. Specifically, the overall AAMRs for these groups are as follows: NH Black: 28.8 (95% CI: 28.0-29.6); Hispanic: 22.8 (95% CI: 22.0-23.6); NH White: 19.5 (95% CI: 19.3-19.7); NH Other: 18.1 (95% CI: 17.1-19.1), all statistically significant with p < 0.001.

Furthermore, between 1999 and 2023, AAMRs across all racial groups have shown a decline, albeit to varying extents, with the most pronounced reduction occurring in Non-Hispanic Others [NH Other: AAPC: -1.51 (CI: -1.86 to -1.11) (p < 0.000001); Hispanic: AAPC: -1.44 (CI: -1.78 to -1.06) (p < 0.000001); NH White: AAPC: -0.74 (CI: -0.92 to -0.58) (p < 0.000001); in contrast, NH Black indicated no significant decline with an AAPC of -0.07 (CI: -0.36 to 0.23) (p = 0.671066) (Supplementary Table 5 and Fig 3).

**Fig. 3:**
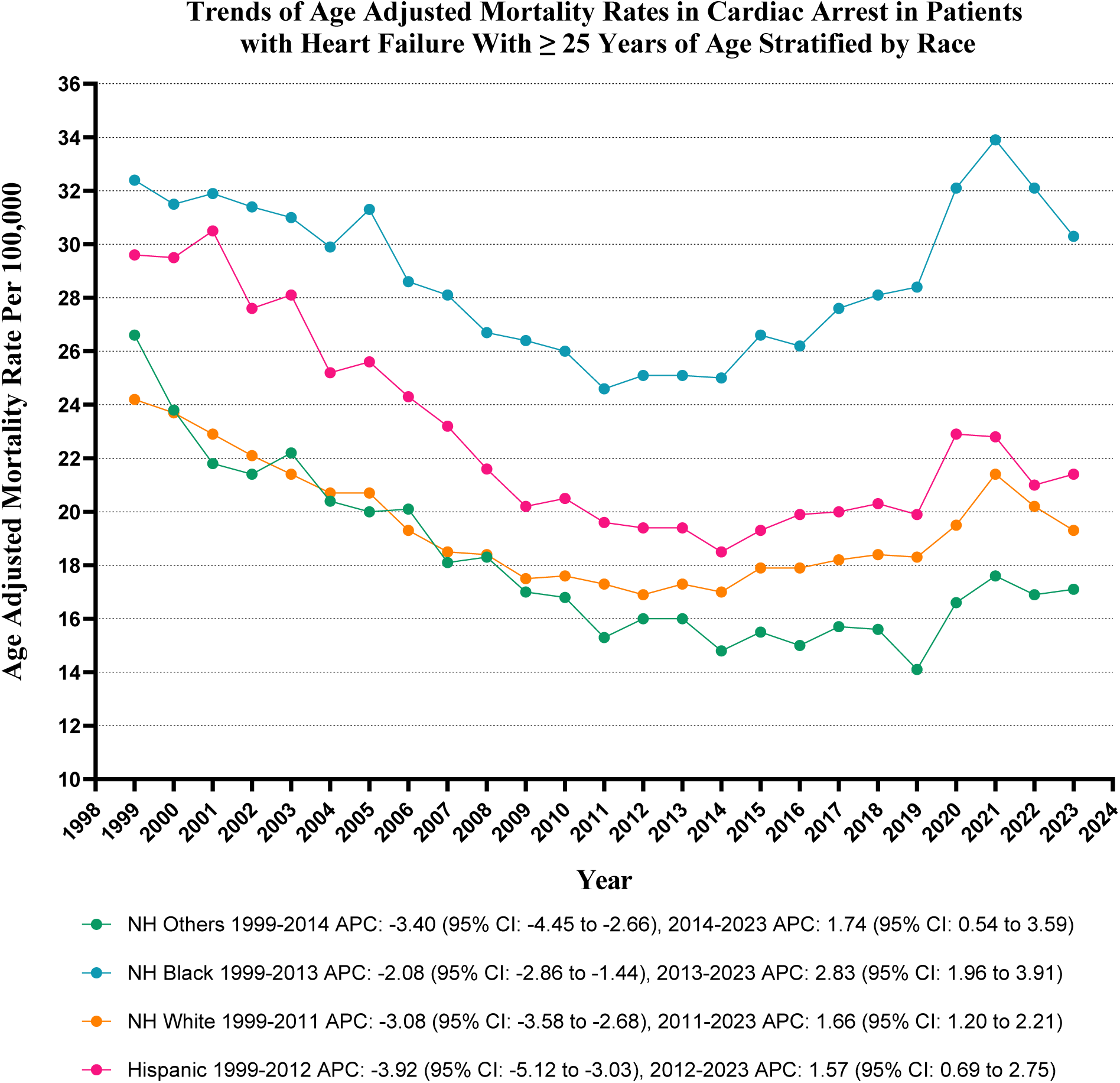
Cardiac Arrest-related age-adjusted mortality rates per 100,000 Stratified by Race in Adults with Heart Failure in the United States, 1999 to 2023

### Cardiac Arrest in Heart Failure-Related AAMR Stratified by Geographical Regions

Notable disparities in AAMRs exist among states, with Maryland reporting a low of 4.3 (95% CI: 4.2-4.5) while Mississippi records a staggering 52.8 (95% CI: 52.1-53.4). The states within the top 90th percentile—including California, Connecticut, Georgia, Mississippi, Nebraska, Nevada, New York, and West Virginia—exhibit AAMRs nearly three times greater than those in the bottom 10th percentile, which comprises Alaska, Delaware, Illinois, Maine, Maryland, Minnesota, Oregon, and Wisconsin (Supplementary Table 6).

Throughout the study, the Western region exhibited the highest AAMR at 28.9 (95% CI: 28.4 to 29.4), followed by the Northeastern region at 25.6 (95% CI: 25.1 to 26.1), the Southern region at 17.9 (95% CI: 17.6 to 18.2), and finally the Midwestern region at 12.5 (95% CI: 12.3 to 12.8). All regional differences are statistically significant, with p < 0.001 (Supplementary Table 7 and Fig 4).

**Fig. 4:**
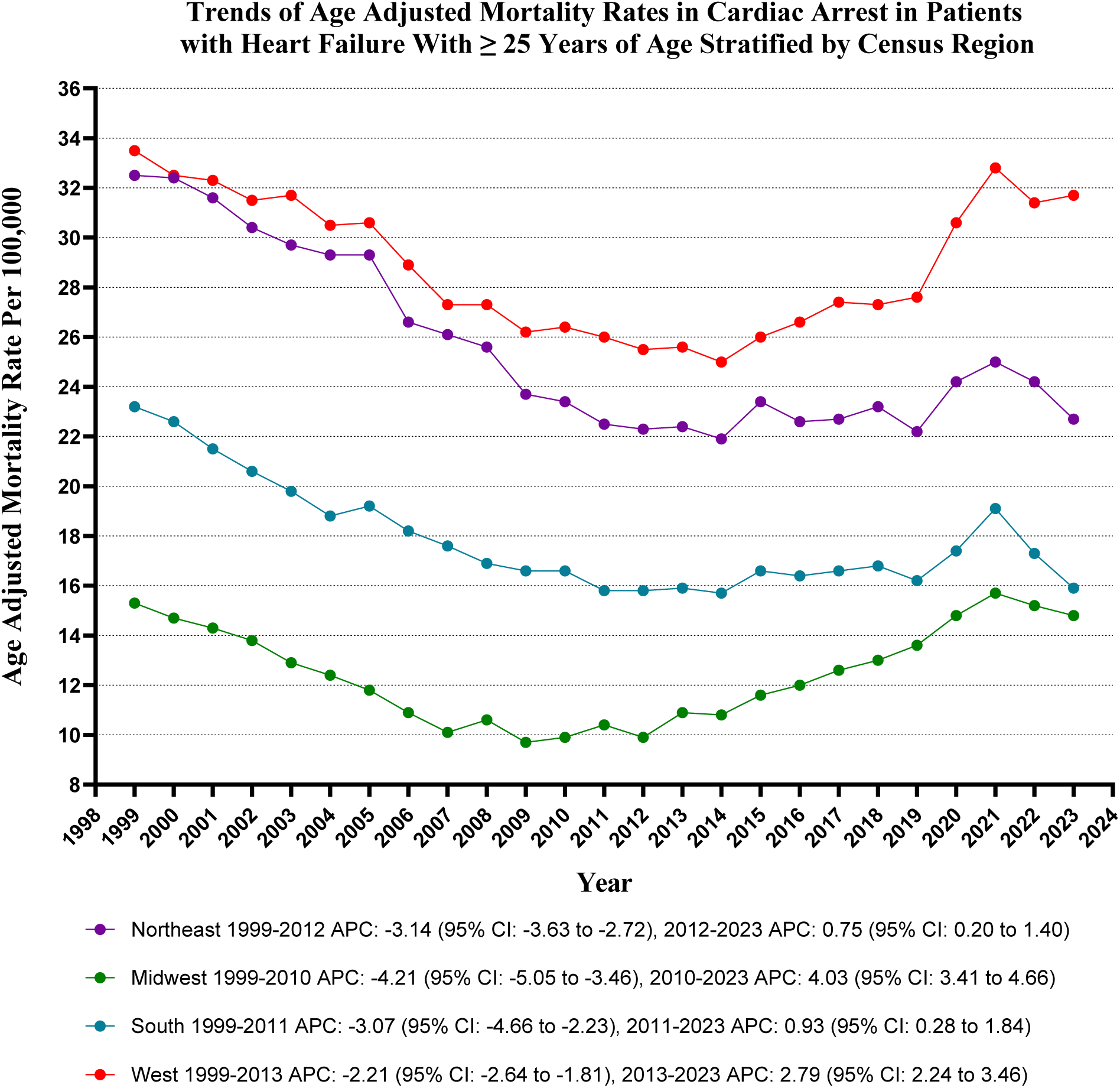
Cardiac Arrest-related age-adjusted mortality rates per 100,000 Stratified by Regions in Adults (≥25 Years) with Heart Failure in the United States, 1999 to 2023

Moreover, it is notable that rural areas have consistently presented with higher AAMRs compared to their urban counterparts throughout most of the study period, reporting overall AAMRs of 20.9 (95% CI: 20.8 to 21.0) versus 20.1 (95% CI: 20.0 to 20.1) for urban areas. While urban and rural AAMRs experienced declines from 1999 to 2023, the decline in urban areas was particularly marked. Specifically, urban areas saw an AAPC of -1.11 (CI: -1.28 to - 0.96) (p < 0.000001), while rural regions reflected a more modest AAPC of -0.51 (CI: -0.69 to - 0.31) (p < 0.000001) (Supplementary Table 8 and Fig 5).

**Fig. 5:**
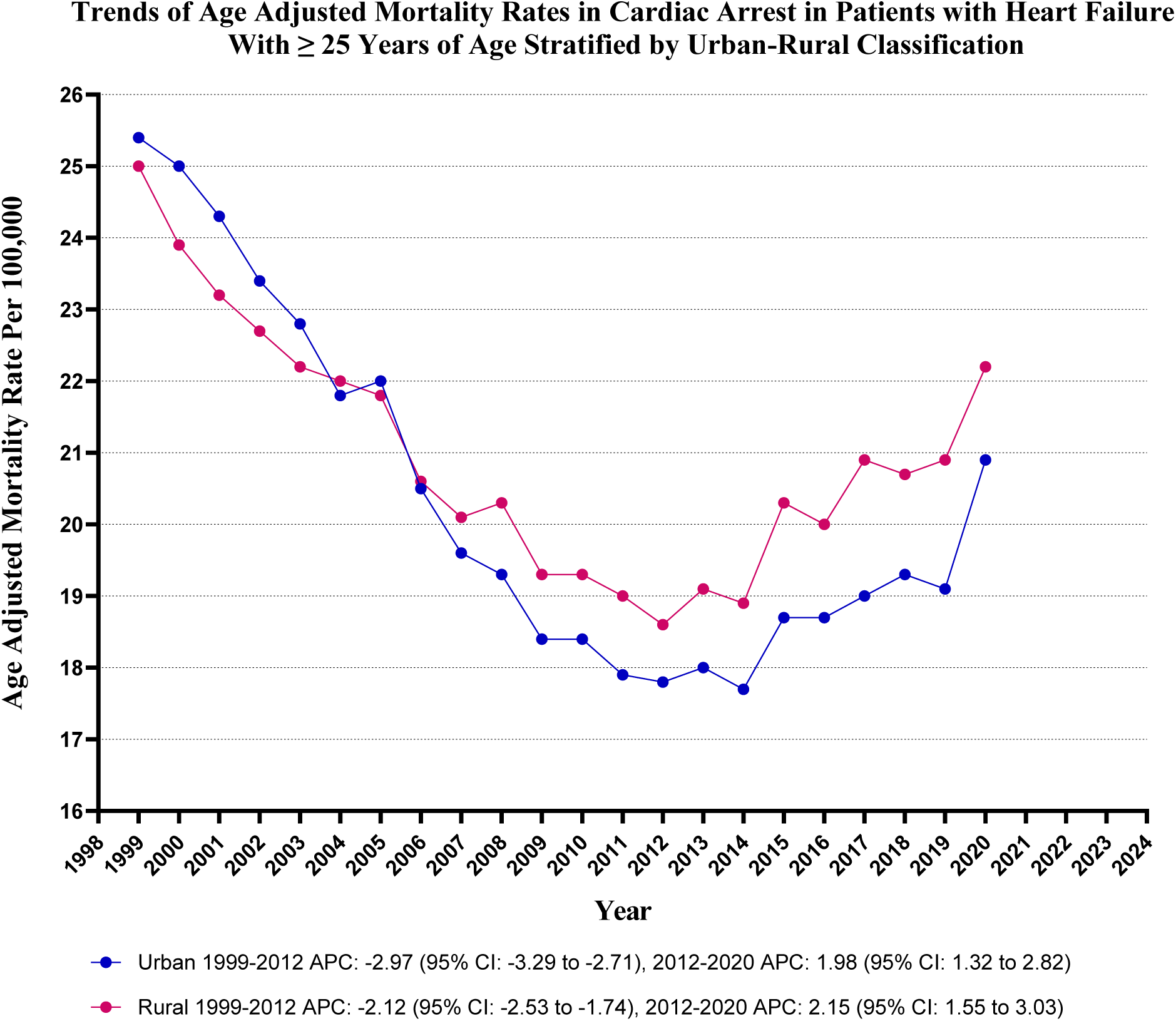
Cardiac Arrest-related age-adjusted mortality rates per 100,000 Stratified by Urbanization in Adults (≥25 Years) with Heart Failure in the United States, 1999 to 2023

## Discussion

Our thorough analysis of mortality data from the CDC WONDER, spanning from 1999 to 2023, highlights critical trends in cardiac arrest mortality among individuals with heart failure. The insights gained from our study are vital for effectively understanding and combating cardiac arrest mortality, and they include the following key findings:

- Cardiac arrest has led to a staggering total of 1,139,963 deaths among adults aged 25 and older with heart failure in the United States.
- The overall age-adjusted mortality rate (AAMR) for cardiac arrest related to heart failure among adults has shown a steady yet significant decline, from 25.3 in 1999 to 20.6 per 100,000 in 2023. Notably, there was a notable surge in mortality between 2018 and 2021, with an annual percentage change (APC) of 5.10. This trend was reversed from 2021 to 2023 with a significant decline with an APC of -4.05
- Men exhibited significantly higher AAMRs compared to adult women, with rates of 24.6 and 17.6, respectively. A dramatic increase in AAMRs was observed between 2018 and 2021, particularly among men, who experienced an annual percentage change (APC) of 5.82.
- Moreover, our analysis revealed that Black individuals exhibited higher AAMRs compared to Hispanic individuals. Notably, Black individuals did not demonstrate a decline in annual percentage changes (APC) during the study period, unlike other demographic groups, which exhibited a consistent downward trend.
- Our analysis reveals significant disparities in age-adjusted mortality rates (AAMRs) across different states. Mississippi exhibited a markedly elevated rate of 52.8, in contrast to Maryland, which reports a significantly lower AAMR of 4.3.

Our analysis shows a notable decline in AAMR from 1999 to 2011 (APC: -2.95). This aligns with earlier research that found a significant drop in cardiovascular deaths (18). This decline primarily resulted from adopting evidence-based therapies, improved heart failure management, and effective public health initiatives targeting cardiovascular risk factors (19,20). Advances in secondary prevention after acute coronary events and better revascularization techniques also played a role (21). In stark contrast, the stagnation in mortality rates observed from 2012 to 2018 (APC: 1.13) raises critical concerns (22). This period suggests that the increasing prevalence of comorbidities, especially diabetes and obesity, along with an aging population, may have counteracted the therapeutic gains achieved in the preceding years (23).

Furthermore, the significant uptick in AAMR from 2018 to 2021 (APC: 5.10) correlates with the emergence of the COVID-19 pandemic, highlighting compelling evidence that cardiovascular mortality escalated during this time due to barriers in healthcare access, disruptions in chronic disease management, and the direct cardiovascular implications of SARS-CoV-2 infection (24,25). Notable is the subsequent reversal in this trend from 2021 to 2023 (APC: -4.05), which aligns with post-pandemic recovery patterns, illustrating how enhanced healthcare access and targeted initiatives aimed at reducing cardiovascular risks contributed to a decline in mortality rates. These significant findings underscore the profound impacts of the pandemic on cardiovascular outcomes and highlight the urgent necessity for healthcare systems to cultivate adaptability and resilience to navigate future global health challenges effectively (20).

Our analysis has uncovered compelling trends in AAMRs between men and women, revealing that men consistently exhibit higher rates throughout the observed period. This is concerning, as men are more prone to cardiovascular diseases, including heart failure (26). Contributing factors include biological differences and lifestyle choices. At the same time, women face unique challenges such as breast cancer therapy, stress-induced cardiomyopathy, a higher prevalence of autoimmune diseases, and the physiological changes linked to pregnancy—which further complicate their cardiovascular health (27).

The rise in cardiac arrest mortality from 2018 to 2021 is likely linked to the COVID-19 pandemic, which exacerbated cardiovascular issues. Despite this, men still show higher AAMRs than women, echoing previous findings (28). Encouragingly, the noticeable decline in mortality rates among women suggests that targeted interventions—such as awareness campaigns and tailored treatment protocols—are making a significant impact in addressing the specific needs of female patients (29).

Our research underscores the troubling racial and ethnic disparities in cardiac arrest mortality, with non-Hispanic Black individuals suffering the highest AAMR of 28.8 per 100,000. This finding is corroborated by the previous study done by Becker et al. under The CPR Chicago Project, which show that socioeconomic factors, including inadequate insurance and limited healthcare access, contribute to these disparities (30,31). Additionally, delays in treatment and a higher prevalence of underlying health conditions lead to increased mortality rates for Black individuals (32). The decline in AAMR among Hispanic and Non-Hispanic Other groups, juxtaposed with the stagnation in Non-Hispanic Black communities, highlights the urgent need for targeted public health initiatives.

Geographically, Mississippi has the highest AAMR at 52.8 per 100,000, a trend attributed to a high prevalence of cardiovascular risk factors and access issues (21,23). The higher mortality rates in rural areas (20.9 per 100,000) compared to urban settings (20.1 per 100,000) further highlight the necessity for improved healthcare access and resources in these communities, as rural populations often face more significant barriers to receiving timely medical care (33). These findings emphasize the need for targeted regional interventions to address modifiable risk factors and enhance emergency cardiac care infrastructure.

## Limitations

This study has limitations, primarily stemming from its retrospective design. The reliance on death certificates sourced from the CDC WONDER database raises concerns about the potential for inaccurate diagnoses, which could result in misclassification bias. Additionally, the lack of laboratory values, medication lists, and clinical information regarding overall health, comorbidities, and treatment restricts a comprehensive understanding of mortality patterns. However, in contrast to existing literature, this study examines adults aged 25 and older across all racial backgrounds, thereby providing an in-depth analysis of cardiac arrest-related mortality trends within a diverse population. From 1999 to 2023, our research presents a long-term perspective that significantly contributes to understanding cardiac arrest mortality’s evolution over more than two decades.

## Conclusion

The analysis identifies significant demographic and geographic disparities in mortality rates. The rising mortality rates associated with cardiac arrest in heart failure patients, particularly among men, Black individuals, and residents of nonmetropolitan areas, necessitate urgent attention and targeted public health interventions to address these disparities. To effectively alleviate these issues and improve health outcomes for this population, it is crucial to implement specific interventions and ensure equitable access to healthcare services.

## Author Contributions

ANM and SN led the study and contributed to the analysis and writing of the original manuscript. SD and GCF supervised the project and helped with editing. The rest of the authors assisted in writing the manuscript.

## Data Availability

Data was available in the manuscript and supplemental files.

## Acknowledgments

The authors acknowledge using the CDC WONDER database to provide the data utilized in this study.

